# Combination therapy of Tocilizumab and steroid for management of COVID-19 associated cytokine release syndrome: A single center experience from Pune, Western India

**DOI:** 10.1101/2021.02.04.21249959

**Authors:** Ameet Dravid, Reema Kashiva, Zafer Khan, Danish Memon, Aparna Kodre, Prashant Potdar, Milind Mane, Rakesh Borse, Vishal Pawar, Dattatraya Patil, Debashis Banerjee, Kailas Bhoite, Reshma Pharande, Suraj Kalyani, Prathamesh Raut, Madhura Bapte, Anshul Mehta, M Sateesh Reddy, Krushnadas Bhayani, S S Laxmi, P D Vishnu, Shipra Srivastava, Shubham Khandelwal, Sailee More, Rohit Shinde, Mohit Pawar, Amol Harshe, Sagar Kadam, Uma Mahajan, Gaurav Joshi, Dilip Mane

**Affiliations:** Department of Infectious diseases and HIV/AIDS, Noble hospitals and Research Centre, Pune, Maharashtra, India; Department of Medicine, Noble hospital and Research Centre, Pune, Maharashtra, India; Department of Critical Care Medicine, Noble hospital and Research Centre, Pune, Maharashtra, India; Department of Pathology, Noble hospital and Research Centre, Pune, Maharashtra, India; Department of Radiology, Noble hospital and Research Centre, Pune, Maharashtra, India; Statistician, VMK Diagnostics private limited, Pune, Maharashtra, India; Independent statistical consultant, Chicago, USA

## Abstract

**Background:** Cytokine release syndrome (CRS) or cytokine storm is thought to be the cause of inflammatory lung damage, worsening pneumonia and death in patients with COVID-19. Steroids (Methylprednisolone or Dexamethasone) and Tocilizumab (TCZ), an interleukin-6 receptor antagonist, are approved for the treatment of CRS in India. The aim of this study was to evaluate the efficacy and safety of combination therapy of TCZ and steroids in COVID-19 associated CRS.

**Methods:** This retrospective cohort study was conducted at a tertiary level private hospital in Pune, India between 2^nd^ April and 2^nd^ November 2020. All patients administered TCZ and steroids for treatment of CRS were included. The primary endpoint was incidence of all-cause mortality. Secondary outcomes studied were need for mechanical ventilation and incidence of infectious complications. Baseline and time-dependent risk factors significantly associated with death were identified by Relative risk estimation.

**Results:** Out of 2831 admitted patients, 515 (24.3% females) were administered TCZ and steroids. Median age of the cohort was 57 (IQR: 46.5, 66) years. Almost 72 % patients had preexisting co-morbidities. Median time to TCZ administration since onset of symptoms was 9 days (IQR: 7, 11). 63% patients needed intensive care unit (ICU) admission. Mechanical ventilation was required in 242 (47%) patients. Of these, 44.2% (107/242) recovered and were weaned off the ventilator. There were 135 deaths (26.2%), while 380 patients (73.8%) had clinical improvement. Infectious complications like hospital acquired pneumonia, bloodstream bacterial and fungal infections were observed in 2.13 %, 2.13 % and 0.06 % patients respectively. Age ≥ 60 years (p=0.014), presence of co-morbidities like hypertension (p = 0.011), IL-6 ≥ 100 pg/ml (p = 0.002), D-dimer ≥ 1000 ng/ml (p < 0.0001), CT severity index ≥ 18 (p < 0.0001) and systemic complications like lung fibrosis (p = 0.019), cardiac arrhythmia (p < 0.0001), hypotension (p < 0.0001) and encephalopathy (p < 0.0001) were associated with increased risk of death.

**Conclusions:** Combination therapy of TCZ and Steroids is likely to be safe and effective in the management of COVID-19 associated cytokine release syndrome. Efficacy of this anti-inflammatory combination therapy needs to be validated in randomized controlled clinical trials.

## Introduction

In December, 2019, Wuhan city, the capital of Hubei province in China, became the centre of an outbreak of viral pneumonia. By Jan 7, 2020, scientists had isolated a novel RNA, beta coronavirus from these patients. It was named severe acute respiratory syndrome coronavirus 2 (SARS-CoV-2) due to its sequence homology with SARS-COV-1 ^[1]^. The disease caused by SARS-COV-2 was later designated coronavirus disease 2019 (COVID-19) in February 2020, by World health organization (WHO) ^[2]^. COVID-19 spread rapidly worldwide and India was no exception. By 2^nd^ November 2020 there have been more than 9 million infections and 0.14 million deaths due to COVID-19 in India ^[3]^.

The clinical presentation of COVID-19 is highly heterogeneous, ranging from asymptomatic cases, mild infection and severe pneumonia. In severe and critical cases, which occur in approximately 15 % and 5 % of patients, pneumonia can lead to acute respiratory distress syndrome (ARDS) that could need invasive mechanical ventilation ^[1, 4-7]^. Critically ill COVID-19 patients have a mortality rate ranging from 35 to 62% ^[8-10]^.

The disease is characterized by 2 phases; viral replication phase and the host inflammatory response phase ^[11]^. Host inflammatory response phase leading to inflammatory lung damage is usually seen 7 days after symptom onset. SARS-CoV-2 can replicate within the pulmonary tissue, activate innate immune response, leading to production of cytokines (Interleukin-1 beta (IL-1B), Interleukin-6 (IL-6) and Tumor necrosis factor (TNF)) by alveolar macrophages which are required for recruitment of adaptive immunity cells. The transition between innate and adaptive immune responses is critical for the clinical trajectory of SARS-CoV-2 infection ^[11-15]^. Adaptive immune response controlled by immune regulatory cells can be a protective immune response or a dysregulated and exacerbated inflammatory response ^[12]^. The protective response is T cell dependent, with CD4 cells helping B cells to produce specific neutralizing antibodies against viral spike (S) protein and cytotoxic CD8 cells eliminating virus infected cells. Protective immune response is present in patients with asymptomatic, mild and some moderate infections that do not progress to severe disease ^[12]^. However, amongst severe and critically ill COVID-19 patients, there is a dysregulated pulmonary and systemic immune response. This dysregulated immune response also known as cytokine release syndrome (CRS) or cytokine storm is characterized by activation of innate immune system, elevation in systemic inflammatory markers (C-reactive protein (CRP), ferritin, lactate dehydrogenase (LDH) and D-dimer) and aberrant pro-inflammatory cytokine secretion (IL-6, soluble IL-2 receptor [IL-2R], IL-10, TNF-α) by alveolar macrophages. It is also accompanied by depleted adaptive immune response. Lymphopenia (decline in CD4+ T cell, CD8+ T cell, Natural killer cell but not in B cell subset) and decreased Interferon Gamma (IFN-γ) expression in CD4+ T cells (CD4+ T cell dysfunction) are immediate consequences of decline in adaptive immunity ^[12-15]^. Cytokine storm leads to destruction of alveolar epithelial cells, increased pulmonary vascular permeability, worsening pneumonia, worsening oxygenation, increased risk of thrombosis and progression to acute respiratory distress syndrome (ARDS) [15]. Rising levels of interleukin-6 (IL-6) in severe COVID-19 have been associated with increased likelihood of ARDS, mechanical ventilation and mortality ^[16-19]^.

Steroids, namely Dexamethasone and Methylprednisolone have been extensively used to resolve hyperinflammation and inflammatory lung damage in COVID-19 ^[20-23]^. After the publication of RECOVERY trial, Dexamethasone was approved by World health organization (WHO) as an immunomodulatory drug for use in hospitalized COVID-19 patients who require oxygen. The benefit of Dexamethasone was greatest for patients who were receiving invasive mechanical ventilation at the time of randomization ^[21]^. In patients with moderate to severe COVID-19, an early short course of methylprednisolone had a reduced rate of the primary composite endpoint of death, ICU transfer, and mechanical ventilation ^[22]^. In patients already on mechanical ventilation, use of Methylprednisolone was associated with increased ventilator-free days and higher probability of extubation ^[23]^. However steroids alone might not be able to tackle cytokine storm in all patients. Other Immunomodulatory drugs such as selective cytokine inhibitors could be of incremental benefit to suppress the hyperinflammation if used in combination with steroids.

Tocilizumab (TCZ) is a recombinant humanized monoclonal interleukin-6 receptor (IL-6R) antibody of the IgG1 subtype. TCZ specifically binds and inhibits soluble and membrane-bound IL-6 receptors (sIL-6R and mIL-6R) and terminates downstream intracellular signal transduction ^[24, 25]^. It has been approved for the treatment of rheumatoid arthritis ^[26]^ and systemic juvenile idiopathic arthritis ^[27]^. In addition, it has also been used in treatment of Castleman disease ^[28]^ and Crohn’s disease ^[29]^. In August 2017, the United States Food and Drug Administration (FDA) approved TCZ for the treatment of cytokine release syndrome (CRS) caused by chimeric antigen receptor T-cell (CAR-T) immunotherapy ^[30]^. Indian council of Medical research (ICMR) guidelines published by Government of India for guidance of physicians in clinical management of COVID-19, have included TCZ as investigational therapy for off label use in patients with moderate disease with progressively increasing oxygen requirements and in mechanically ventilated patients not improving despite use of steroids ^[31]^.

This recommendation stems from multiple retrospective and small prospective studies ^[32-58]^, that have suggested strong benefits due to use of TCZ in form of reduced risk of invasive mechanical ventilation or death in patients with severe COVID-19. On the other hand, data from randomized controlled trials (RCT) has shown disappointing results with evidence of modest efficacy and no mortality benefit ^[59-63]^. RCT’s have shown that TCZ reduces need for ICU admission and mechanical ventilation in patients with severe COVID-19 ^[61, 62]^. However, out of the 4 RCT’s, only the EMPACTA trial ^[62]^ used concomitant TCZ plus steroids in management of COVID-19 induced hyperinflammation. As a result more evidence regarding positioning of TCZ as an immunomodulatory therapy in COVID-19 needs to be published. Data from resource limited settings like India regarding use of TCZ and steroids in treatment of CRS has also been scarce ^[64, 65]^. The aim of this single center retrospective cohort study conducted in Pune, India was to estimate the efficacy and safety of combination therapy of TCZ and steroid in management of COVID-19 associated CRS.

## Methods

### Study Setting

This retrospective cohort study was conducted at Noble hospital and Research Centre (NHRC), Pune, Western India. NHRC is a tertiary level private hospital designated for clinical management of COVID-19 patients in Pune since 23^rd^ March 2020. Pune is located in the state of Maharashtra, Western India and was one of the epicenters of COVID-19 epidemic in India. As of 2^nd^ November 2020, Maharashtra state has reported more than 1.71 million cases of COVID-19 and more than 45,000 deaths ^[3]^. Till 2^nd^ November 2020, NHRC has admitted 2831 COVID-19 patients.

NHRC provides clinical care, diagnostic and treatment services to COVID-19 patients at a subsidized cost. Patients are referred from primary care physicians, private practitioners of alternative medical systems, primary level COVID care centers (both government run and private owned) and other tertiary level hospitals. Data of all hospitalized COVID-19 patients is entered into an electronic database (Lifeline electronic database, Manorama infosystems, Kolhapur, India). Data was obtained from electronic health record of each individual by manual abstraction. It included hospitalization dates, demographics, co-morbidities, clinical examination data, laboratory data (including inflammatory markers), microbiology reports, imaging reports (X ray chest and High resolution computerized tomography scan (HRCT chest)), data on use of supplemental oxygen, ventilation parameters (noninvasive or invasive ventilation) and hospital outcomes. Ordinal scale for COVID-19 severity was noted for all patients at hospital admission, during hospital stay and at discharge or death. The 8 level ordinal scale is as follows: 1 = ambulatory and no restriction of activities; 2 = ambulatory and restriction of activities due to use of home oxygen therapy or complications; 3 = Hospitalized but no oxygen therapy; 4 = Hospitalized with oxygen therapy by nasal prongs; 5 = Hospitalized with oxygen therapy by Non Re-breathing mask; 6 = Hospitalized with severe disease and on high flow nasal oxygen (HFNO) or Noninvasive ventilation; 7 = Hospitalized with severe disease and on invasive mechanical ventilation; 8 = Death.

### Study Population

Patients were eligible for inclusion in this analysis if they were admitted between 2^nd^ April 2020 and 2^nd^ November 2020 and were administered TCZ. Criteria for prescribing TCZ were developed by the Department of Infectious Diseases and Department of Critical care medicine. Patients were administered TCZ if they satisfied following criteria for hyperinflammation or CRS:

1) Reverse-transcriptase polymerase chain reaction (RT-PCR) test positive for SARS-CoV-2 RNA or positive COVID antibody test.
2) Lung Imaging: Moderate or severe pneumonia on High resolution Computerized tomography scan (HRCT) of chest (CT severity index ≥ 8 ^[66]^) or X ray chest showing evidence of pneumonia.
3) Day 7 to 14 since onset of symptoms.
4a) Rapidly worsening respiratory status despite use of steroids and antiviral drugs: Hypoxia (room air oxygen saturation (SPO2) < 90 %) and tachypnea (respiratory rate > 30 per minute) at rest or after minimal exertion which requires supplemental oxygen. OR
4b) Rapidly worsening respiratory status despite use of steroids and antiviral drugs: Requirement of noninvasive or invasive ventilation to resolve hypoxia and tachypnea. OR
4c) PaO2/FiO2 ratio of less than 300 mm Hg on room air.
5) Elevated inflammatory markers: IL-6 (> 100 pg/ml or 5 fold increase from prior level) or one out of D-dimer (> 1000 ng/ml), Ferritin (> 1000 ng/ml) and CRP (> 10 mg/ml) being elevated.

Exclusion criteria: Following patients were not prescribed TCZ during hospital admission or were excluded from the analysis.

1. Pregnant females.
2. Patients having active systemic infections (bacterial/fungal), active tuberculosis and active hepatitis B/C co-infection.
3. History of diverticulitis or inflammatory bowel disease.
4. Patients who refused TCZ therapy for management of CRS.
5. Patients administered TCZ in another institute prior to transfer to NHRC.

Tocilizumab was given intravenously at 8 mg/kg bodyweight (up to a maximum of 800 mg in two infusions, 12 hours apart). Additional doses were administered if patients were morbidly obese (body weight > 100 kg) or to treat persistent hyperinflammation and worsening ARDS. Patients or their immediate family members signed an informed consent form prior to TCZ administration. The language in the consent form was non-prescriptive, saying that TCZ can be used off-label in patients with COVID-19 induced hyperinflammation as per ICMR guidelines. The consent form cautions that the evidence for benefit is modest; a risk for infectious complications exists but in view of limited treatment options in patients with severe pneumonia, therapy can be considered. The cost of TCZ (approximately 500 dollars for 1 vial of 400 mg) was borne by the patient as an out of pocket expense or by third party reimbursement via medical health insurance.

For all patients administered TCZ, we also scrutinized inpatient case files until hospital discharge, death, or December 2^nd^ 2020—the date on which the database was locked—whichever happened first. Number of TCZ doses given, type of concomitant steroid and dose of steroid given and other concomitant medication prescribed to patient were noted. Time to TCZ administration since onset of symptoms and time to TCZ administration since admission in hospital was calculated. Oxygenation and ventilation outcomes in patients administered TCZ plus steroids were noted. Systemic complications (including infectious complications) developing in patients during hospital admission were also noted. All patients who died during hospital admission were identified and a death audit to look for complications and cause of death was undertaken. All patients who showed clinical improvement, got discharged from hospital and had outpatient follow-up at 15 and 30 days after discharge were identified. Their outpatient follow-up visits were traced from electronic database to look for delayed complications. There was no control group in our study as all patients with suspicion of hyperinflammation or CRS ended up getting TCZ and steroid.

### Concomitant medications given for COVID-19 therapy

NHRC follows the ICMR guidelines published by Government of India for COVID-19 management ^[31]^. These guidelines get updated from time to time. All hypoxic COVID-19 patients who received TCZ in our cohort were already receiving antiviral agents, intravenous steroids (Dexamethasone 6 mg per day or Methylprednisolone 40 mg twice a day) and systemic anticoagulation (Low molecular weight heparin (Enoxaparin) or Unfractionated Heparin) as a standard of care. Intravenous steroids were continued for a maximum of 10 days followed by shift to oral Prednisolone in tapering doses over next 10 days. Hydroxychloroquine and Lopinavir/ritonavir were initially recommended as standard antiviral therapy. Once clinical trial and observational study data about lack of efficacy and toxicity was published, their use as antivirals was discontinued ^[67-69]^. Remdesvir was used as antiviral of choice at our institute since July 26^th^, 2020 and was administered to all patients presenting with moderate or severe pneumonia ^[70]^. Convalescent plasma therapy (CPT) was also used in a subset of patients presenting within 7 days of symptom onset as an antiviral agent. Adjunctive antibiotic and antifungal therapy in patients to prevent bacterial and fungal super-infections was decided by the infectious disease physician. After administration of TCZ, anti-fibrotic agents like Pirfenidone (daily dosage ranging from 800 to 2000 mg per day) and Nintedanib (daily dosage of 150 mg twice a day) were added to the treatment protocol for patients suspected of developing lung fibrosis.

### Outcomes

#### Primary endpoint

1) Deaths in the cohort after TCZ and steroid administration.

#### Secondary outcomes

2) Number of patients who received noninvasive or invasive ventilation: Patients who required mechanical ventilation (noninvasive or invasive) in our cohort were identified. NHRC adopted the delayed intubation and delayed invasive mechanical ventilation (IMV) policy for patients with COVID-19 associated ARDS. Patients not maintaining arterial oxygen saturation (SPO2) > 90 % on supplemental oxygen (Non re-breathing mask at 15 liters/minute) were initially started on noninvasive ventilation (NIV: High flow nasal oxygen (HFNO) or Bi-level positive airway pressure (BIPAP)). The indications for IMV in our hospital was respiratory failure on NIV, defined by any of the following criteria: a) respiratory rate of 40 or more breaths per minute on NIV; b) respiratory distress with activation of accessory respiratory muscles; c) the need for fractional inspired oxygen (FiO2) of 100 % on NIV to maintain an SPO2 level of 90%, or a PaO2/FiO2 ratio of less than 100 mm Hg; d) Neurological deterioration (altered consciousness with a Glasgow Coma Scale score of 10 or higher) on treatment. For those individuals in whom consent for the use of IMV was declined, NIV was continued till either clinical recovery or death.
3) Number of patients who could be weaned off NIV or IMV and discharged from hospital.
4) Improvement in 8 point ordinal scale reflecting declining disease severity.
5) Incidence of infectious complications (bacterial or fungal super-infections) in patients after TCZ and steroid administration: Patients developing fever (Temperature > 38.3 degrees Celsius), productive cough, leucocytosis (WBC count > 15,000 cells/mm^3^ with predominance of Neutrophils and shift to left) or increased serum procalcitonin despite adequate antibiotic and antifungal therapy were investigated for super-infections. All positive blood and respiratory cultures (bacterial and fungal culture) were assessed by an Infectious diseases physician to decide infection versus colonization. Infections were included if they occurred > 48 hours after TCZ administration.
6) Incidence of systemic complications like cardiovascular (acute myocardial infarction, congestive cardiac failure, arrhythmia and myocarditis), respiratory (lung fibrosis and pulmonary embolism), neurologic (stroke, encephalopathy, meningo-encephalitis, transverse myelitis and Guillaine Barre syndrome (GBS)), gastrointestinal (intestinal perforation, gastroenteritis and abdominal blood vessel thrombosis), hepatic (hepatitis) and renal complications (acute kidney injury) was estimated. Hepatitis was defined as increase in total bilirubin or serum glutamate oxaloacetate/pyruvate transferase (SGOT/SGPT) value more than three times above baseline value. Acute kidney injury (AKI) was defined as abrupt (within 48 hours) reduction of kidney function manifesting as a percentage increase in serum creatinine of 50 % or greater (1.5-fold from baseline) or a reduction in urine output, defined as less than 0.5 ml/kg/hour for more than 6 hours ^[71]^. Encephalopathy was defined as change in personality, behavior, cognition, or consciousness (including clinical presentations of delirium or coma) in patients without evidence of brain inflammation (increased cerebrospinal fluid (CSF) protein and CSF pleocytosis) ^[72]^.

The use of database for clinical research was approved by the institutional review board (IRB) of Noble hospital and Research Centre, Pune, India.

## Statistical Methods

Continuous variables were summarized using median and interquartile range (IQR), while categorical variables were summarized using frequency and percentages. Continuous variables were compared using a Mann Whitney U test. Categorical variables were compared using Chi-square test, Proportion test and Fishers’ exact test. Baseline and time-dependent risk factors significantly associated with death were identified by Relative risk estimation. Baseline risk factors included were age (< 60 years or ≥ 60 years), gender, co-morbidities like diabetes, hypertension, ischemic heart disease and chronic kidney disease and baseline investigations like IL-6 (≥ 100 versus < 100 pg/ml), absolute lymphocyte count (< 1000 versus ≥ 1000 cells/mm^3^), D-dimer (≥ 1000 versus < 1000 ng/ml) and CT severity index (≥ 18 versus < 18). Time-dependent risk factors included systemic complications like lung fibrosis, arrhythmia, hypotension, thrombocytopenia, hepatitis, acute kidney injury and encephalopathy. The p value ≤ 0.05 was considered as statistically significant. All data was analyzed by SPSS version 12.0.

## Results

Out of the 2831 COVID-19 patients admitted in NHRC, 515 were administered TCZ. Median age of the cohort was 57 (IQR: 46.5, 66) years and it included 24.3 % females. Two hundred and twenty two (43.1 %) patients were ≥ 60 years of age. Three hundred and seventy three (72.4 %) patients had preexisting co-morbidities, with Diabetes mellitus (45.4 %), Hypertension (48.3 %), Ischemic heart disease (13.6 %), Chronic kidney disease (7.8 %) and Obesity (Body mass index > 30 kg/m^2^, 9.5 %) being the commonest (Table 1). Preexisting co-morbidities, presenting symptoms in patients at admission and investigations performed prior to TCZ therapy are enumerated in Table 1 and 2 respectively. Fever (81%), dry cough (77%), dyspnea on exertion (81%) and bodyache or myalgia (49 %) were the commonest symptoms seen in patients (Table 1). Two hundred and twenty (220/515, 43%) patients had lymphocytopenia (absolute lymphocyte count < 1000 copies/ml) while 16 % (80/515) patients had thrombocytopenia (Platelet count < 150,000 cells/mm^3^) prior to TCZ administration. One hundred and sixty one (161/515, 31.3 %) patients had IL-6 level ≥ 100 pg/ml, 22.7 % (117/515) had CRP ≥ 100 mg/dl, 21 % (108/515) had D-dimer ≥ 1000 ng/ml and 23.7 % (122/515) had serum Ferritin ≥ 1000 mg/dl prior to TCZ administration (Table 2). Out of 373 patients who performed HRCT of Chest (GE Optima, 128 slice CT scanner), CT severity index indicated moderate disease (CT severity index: 8-14) in 37.5 % (140/373) and severe disease (CT severity index: 15-25) in 62.5 % (233/373) of patients. Three hundred and eighty seven (387/515, 75.1%) patients had arterial blood gas analysis performed prior to TCZ therapy. PaO2/FiO2 ratio was in the range of 200-300, 100-200 and < 100 mm Hg amongst 65 (16.8 %), 182 (47 %) and 140 (36.2 %) individuals respectively.

**Table 1:** Baseline characteristics of patients in the cohort.

| **Co-morbidities** | **Total cohort**<br>**n=515 (%)** | **Presenting symptoms** | **n (%)** |
| --- | --- | --- | --- |
| Diabetes mellitus | 234 (45.4%) | Fever | 419 (81%) |
| Hypertension | 249 (48.3%) | Dry cough | 398 (77%) |
| Ischemic heart disease | 70 (13.6%) | Cough with expectoration | 36 (7%) |
| Chronic kidney disease | 40 (7.8%) | Hemoptysis | 12 (2%) |
| Lung disease | 22 (4.3%) | Myalgia or Bodyache | 254 (49%) |
| Liver disease (Liver cirrhosis) | 7 (1.4%) | Chills | 76 (15%) |
| Stroke | 5 (1%) | Headache | 58 (11%) |
| Other Neurologic comorbidities | 10 (1.9%) | Loss of appetite | 73 (14%) |
| History of cancer | 6 (1.1%) | Dyspnoea at rest or on exertion | 419 (81%) |
| Past history of pulmonary Tuberculosis | 5 (1%) | Anosmia | 20 (4%) |
| HIV infection | 3 (0.6%) | Dysgeusia | 59 (11%) |
| Obesity | 49 (9.5%) | Diarrhea | 44 (9%) |
| Thyroid disorder | 41 (7.9%) | Vomiting | 36 (7 %) |
| Rheumatologic disorders | 10 (1.9%) | Abdominal pain | 11 (2%) |
| 0 co-morbidities | 142 (27.6 %) | Chest pain/Chest tightness | 19 (4%) |
| 1 comorbidity | 144 (28 %) | Upper respiratory tract symptoms | 69 (13%) |
| 2 comorbidity | 134 (26 %) | Giddiness | 22 (4%) |
| 3 or more co-morbidities | 95 (18.4 %) |  |  |

**Lung disease:** Chronic obstructive pulmonary disease, Asthma, Interstitial lung disease, Obstructive sleep apnea;
**Other neurologic comorbidities:** Parkinson's disease, Alzheimer's disease, Seizure disorder, Cerebral palsy, Poliomyelitis, Psychiatric disorders; **Rheumatologic co-morbidities:** Rheumatoid arthritis, Gouty arthritis, Psoriasis, Systemic lupus erythematosus, Vasculitis; **Thyroid disease:** Hypothyroidism, Hyperthyroidism;
**Upper respiratory tract symptoms:** Runny nose; Sore throat; Nasal congestion; Sneezing.

**Table 2:** Demographic features and Baseline Investigations in patients administered Tocilizumab and Steroids.

| Investigations | All patients (n=515)<br>Median, IQR | Patients who<br>recovered and were<br>discharged (n= 380),<br>Median, IQR | Patients who<br>died (n= 135),<br>Median, IQR | p value |
| --- | --- | --- | --- | --- |
| Age | 57 (46.5,66) | 55 (45,64) | 62 (53,69) | **< 0.0001** |
| Gender : Male | 390 (75.7%) | 293 (77.1%) | 97 (71.9%) | 0.226 |
| Gender : Female | 125 (24.3%) | 87 (22.9%) | 38 (28.1%) | 0.226 |
| Any co-morbidity | 373 (72.4 %) | 260 (69.7%) | 113 (83.7%) | **0.004** |
| Diabetes mellitus | 234 (45 %) | 167 (44 %) | 67 (49 %) | 0.253 |
| Hypertension | 249 (48 %) | 171 (45 %) | 78 (58 %) | **0.01** |
| Ischemic heart disease | 70 (14 %) | 46 (12 %) | 24 (18 %) | 0.097 |
| Chronic kidney disease | 40 (8 %) | 23 (6 %) | 17 (13 %) | **0.016** |
| Hemoglobin (g/dl) | 13.1 (11.6,14.5) | 13.2 (11.7,14.5) | 12.9 (11.2,14) | 0.092 |
| White blood cell<br>count(cells/mm3) | 6900 (5000,10300) | 6500 (4800,9600) | 8400 (5575,<br>12100) | **< 0.0001** |
| Absolute lymphocyte count<br>(cells/mm3) | 1071 (760,1488) | 1089 (795, 1519) | 1001 (659, 1440) | 0.114 |
| Platelet count<br>(cells/mm3) | 207000<br>(163000, 268000) | 207000<br>(161000, 268500) | 208000<br>(167000, 268000) | 0.477 |
| SGPT (U/L) | 35 (22, 51) | 34 (23,51) | 37 (22,53) | 0.952 |
| SGOT (U/L) | 43 (30,64) | 42 (31,61) | 47(29,73) | 0.219 |
| Blood Urea (mg/dl) | 31 (22,43) | 29 (20,39) | 37 (25,55) | **< 0.0001** |
| Creatinine (mg/dl) | 1.08 (0.93,1.19) | 1.07 (0.93,1.19) | 1.12 (0.94,1.31) | 0.105 |
| IL-6<br>Normal range: 0-7 pg/ml | 69.83<br>(32.53, 117.00) | 60.17<br>(31.28, 105.85) | 87.20<br>(48.00, 161.90) | **< 0.0001** |
| CRP<br>Normal range: 0-6 mg/L | 54.92<br>(30.36, 96.29) | 53.42<br>(30.99 , 85.56) | 62.74<br>(27.29, 110.08) | 0.190 |
| D dimer<br>Normal range: < 252 ng/ml | 344.0<br>(256.0, 814.5) | 318.5<br>(251.0, 568.0) | 593.0<br>(316.0, 1200) | **< 0.0001** |
| Ferritin<br>Normal range: 15-150 ng/ml | 494.45<br>(238.30, 976.80) | 448.50<br>(219.60, 855.10) | 687.40<br>(305.90,1274.00) | **0.002** |
| Procalcitonin<br>Normal range: < 0.05 ng/ml | 0.21 (0.17, 0.28) | 0.21 (0.17, 0.28) | 0.22 (0.17, 0.28) | 0.391 |
| CT severity index | 16 (12,18) | 15 (12,17) | 18 (16, 20) | **< 0.0001** |

SGOT: Serum glutamate ornithyl transferase; SGPT: Serum glutamate pyruvate transferase; IL-6: Interleukin-6; CRP: C reactive protein; CT: Computerized tomography scans.

Total cumulative dose of TCZ administered was 400 mg, 800 mg and 1200 mg in 47.2 % (243/515), 48 % (247/515) and 4.8 % (25/515) patients respectively. Median time to TCZ administration since onset of symptoms was 9 days (IQR: 7, 11) and median time to TCZ administration since hospital admission was 2 days (IQR: 2, 4). Overall 87.2 %, 90 % and 94.4 % patients were prescribed Remdesvir, Methylprednisolone and Low molecular weight or conventional Heparin along with TCZ (Table 3). Intensive care unit (ICU) admission was needed in 63.6 % (327/515) of patients while 36.4 % (187/515) patients recovered in isolation wards. Two hundred and seventy three patients (53 %) required only supplemental oxygen prior to recovery (improvement in ordinal scale from 4 or 5 to 1). Seventy one patients were already on NIV or IMV (HFNO-27, BIPAP-41, IMV-3) prior to TCZ administration while 171 patients needed it after TCZ administration. Overall, 242 patients (47%) needed NIV and/or IMV, of which 44.2 % (107/242) recovered and were weaned off the ventilator (improvement in ordinal scale from 6 or 7 to 1). Two hundred and twenty three (223/242, 92.1%) patients required NIV (HFNO/BIPAP) during the duration of the study out of which 104 (46.6%) could be weaned off NIV and discharged from hospital (improvement in ordinal scale from 6 to 1). One hundred and thirty (130/242, 53.7 %) patients required IMV, of whom only 3 survived (improvement in ordinal scale from 7 to 1). Median time to hospital discharge or death was 13 days (IQR: 10, 17). Among patients admitted in ICU, median duration of ICU stay was 10 days (IQR: 7, 16). In patients needing mechanical ventilation (NIV or IMV), median duration of ventilation was 10 days (IQR: 6, 13). Median time spent on NIV was 7 (IQR: 3, 12) days while median time spent on IMV was 3 (IQR: 2, 6) days. Twenty one (4.1 %) patients were administered hemodialysis while 105 (20.4 %) needed vasopressor agents like Noradrenaline and Vasopressin for hypotension (Table 3).

**Table 3:** Concomitant drugs administered along with Tocilizumab.

| **Concomitant Medication (N=515)** | **N** | **%** |
| --- | --- | --- |
| Remdesvir | 449 | 87% |
| Methylprednisolone | 463 | 90% |
| Dexamethasone | 52 | 10% |
| Enoxaparin/ Conventional Heparin | 486 | 94% |
| Hydroxychloroquine | 16 | 3.1% |
| Lopinavir-ritonavir | 231 | 45% |
| Convalescent plasma therapy | 231 | 45% |
| Methylene blue | 153 | 30% |
| Pirfenidone | 239 | 46.41% |
| Nintedanib | 81 | 15.73% |
| Piperacillin-Tazobactam | 154 | 30% |
| Meropenem | 359 | 70% |
| Teicoplanin | 324 | 63% |
| Fluconazole | 382 | 74.2% |
| Caspofungin | 99 | 19.2% |
| Noradrenaline | 105 | 20.4 % |
| Vasopressin | 41 | 8% |
| Hemodialysis | 21 | 4% |

Systemic complications (fatal and nonfatal) observed in patients in our cohort are enumerated in Table 4. Septic shock (persistent hypotension requiring vasopressors to maintain systolic blood pressure > 100 mm Hg and a serum lactate level greater than 2 mmol/L) was noted in 58 (11.3 %) patients, of whom 4 recovered after treatment. Infectious complications like hospital or ventilator associated pneumonia (HAP/VAP), bloodstream bacterial infections and bloodstream fungal infections were seen in 2.13 % (11/515), 2.13 % (11/515) and 0.06 % (3/515) patients respectively. Methicillin resistant Staphylococcus aureus (MRSA) was the commonest bacteria (6/11 cases) and Candida albicans was the commonest fungus (3/3 cases) isolated in blood culture. Multidrug resistant Gram negative bacilli (Pseudomonas Aeruginosa (3/11), Acinetobacter Baumanii (3/11) and Klebsiella pneumonia (2/11)) were the commonest causative agents of HAP or VAP.

**Table 4:** Systemic complications in patients after administration of Tocilizumab and Steroids.

| Type of complication | Total cohort<br>n =515 (%) | Patients who<br>recovered<br>n = 380 (%) | Patients who<br>died<br>n = 135 (%) | p value |
| --- | --- | --- | --- | --- |
| Subcutaneous emphysema/ Pneumothorax | 26 (5.1 %) | 1 (0.3 %) | 25 (18.5 %) | **<0.0001** |
| Hemoptysis | 23 (4.5 %) | 18 (4.7 %) | 5 (3.7 %) | 0.628 |
| Lung fibrosis | 136 (26.4%) | 90 (23.7 %) | 46 (34.1 %) | **0.019** |
| Arrhythmia | 34 (6.63 %) | 6 (1.6 %) | 28 (20.7 %) | **<0.0001** |
| Congestive cardiac failure | 6 (1.2 %) | 4 (1.02%) | 2 (1.5%) | 0.655 |
| Acute myocardial infarction | 5 (0.97%) | 2 (0.5 %) | 3 (2.2 %) | 0.116 |
| Hypotension | 105 (20.4 %) | 14 (3.7 %) | 91 (67.4 %) | **<0.0001** |
| Thrombocytopenia | 59 (11.5%) | 11 (2.9 %) | 48 (35.6 %) | **<0.0001** |
| Bleeding episodes | 14 (2.7 %) | 6 (1.6 %) | 8 (6 %) | **0.008** |
| Deep vein thrombosis | 1 (0.19%) | 0 | 1 (0.76%) | N/A |
| Pulmonary embolism | 1 (0.19%) | 0 | 1 (0.76%) | N/A |
| Thrombosis of abdominal vessels | 2 (0.4%) | 0 | 2 (1.5%) | N/A |
| Intestinal perforation | 2 (0.4%) | 0 | 2 (1.5%) | N/A |
| HAP/VAP | 11 (2.1%) | 1 (0.3 %) | 10 (7.6 %) | **<0.0001** |
| BSI Bacterial | 11 (2.1 %) | 5 (1.3 %) | 6 (4.6%) | **0.031** |
| BSI Fungal | 3 (0.58%) | 2 (0.5 %) | 1 (0.8 %) | >0.999 |
| Septic shock | 58 (11.3 %) | 4 (1.1 %) | 54 (40 %) | **<0.0001** |
| Encephalopathy | 44 (8.5 %) | 10 (2.6 %) | 34 (25.2 %) | **<0.0001** |
| Stroke | 5 (0.97%) | 1 (0.3%) | 4 (3%) | **0.018** |
| Guillaine Barre syndrome | 1 (0.2%) | 1 (0.3%) | 0 | N/A |
| Acute kidney injury | 43 (8.3 %) | 30 (7.7 %) | 13 (9.8 %) | 0.539 |
| Hepatitis | 53 (10.3 %) | 37 (9.7 %) | 16 (11.4 %) | 0.700 |

Arrhythmia: atrial fibrillation, atrial flutter, ventricular tachycardia, and ventricular fibrillation; HAP/VAP: Hospital associated pneumonia/ Ventilator associated pneumonia; BSI Bacterial: Blood stream bacterial infection; BSI fungal: Bloodstream fungal infection. N/A: Not applicable. p value in bold indicates statistically significant value.

There were 135 deaths (26.2 %) during hospital admission. ICU mortality rate was 40.4% while mortality rate amongst patients requiring NIV/IMV was 55.8 %. Of the 71 patients already on NIV or IMV prior to TCZ administration, 70.4 % (50/71) died while 29.6 % (21/71) survived and were discharged from hospital. Out of the 171 patients who required NIV and/or IMV after TCZ administration, 49.7 % (85/171) died while 50.3 % (86/171) recovered. Commonest cause of death was acute respiratory distress syndrome leading to respiratory failure. The relative risk (RR) of death was significantly higher in patients with age ≥ 60 years (RR=1.555 (95% CI: 1.283, 1.884)), concomitant hypertension (RR=1.284 (95% CI: 1.070, 1.540)), preexisting chronic kidney disease (RR=2.081 (95% CI: 1.147, 3.773)), IL-6 ≥ 100 pg/ml (RR=1.515 (95% CI: 1.172, 1.956)), D-dimer ≥ 1000 ng/ml (RR=2.134 (95% CI: 1.542, 2.954)) and CT severity index ≥ 18 (RR=2.491 (95% CI: 1.882, 3.299)). Development of systemic complications like lung fibrosis (RR=1.439 (95% CI: 1.07, 1.934)), cardiac arrhythmia (RR=13.136 (95% CI: 5.561, 31.029)), hypotension (RR=18.296 (95% CI: 10.798, 31.000)), new onset or worsening thrombocytopenia (RR=12.283 (95% CI: 6.574, 22.949)) and encephalopathy (RR=9.570 (95% CI: 4.862, 18.837)) were also associated with increased risk of death (Figure 1, Table 5).

**Table 5:**
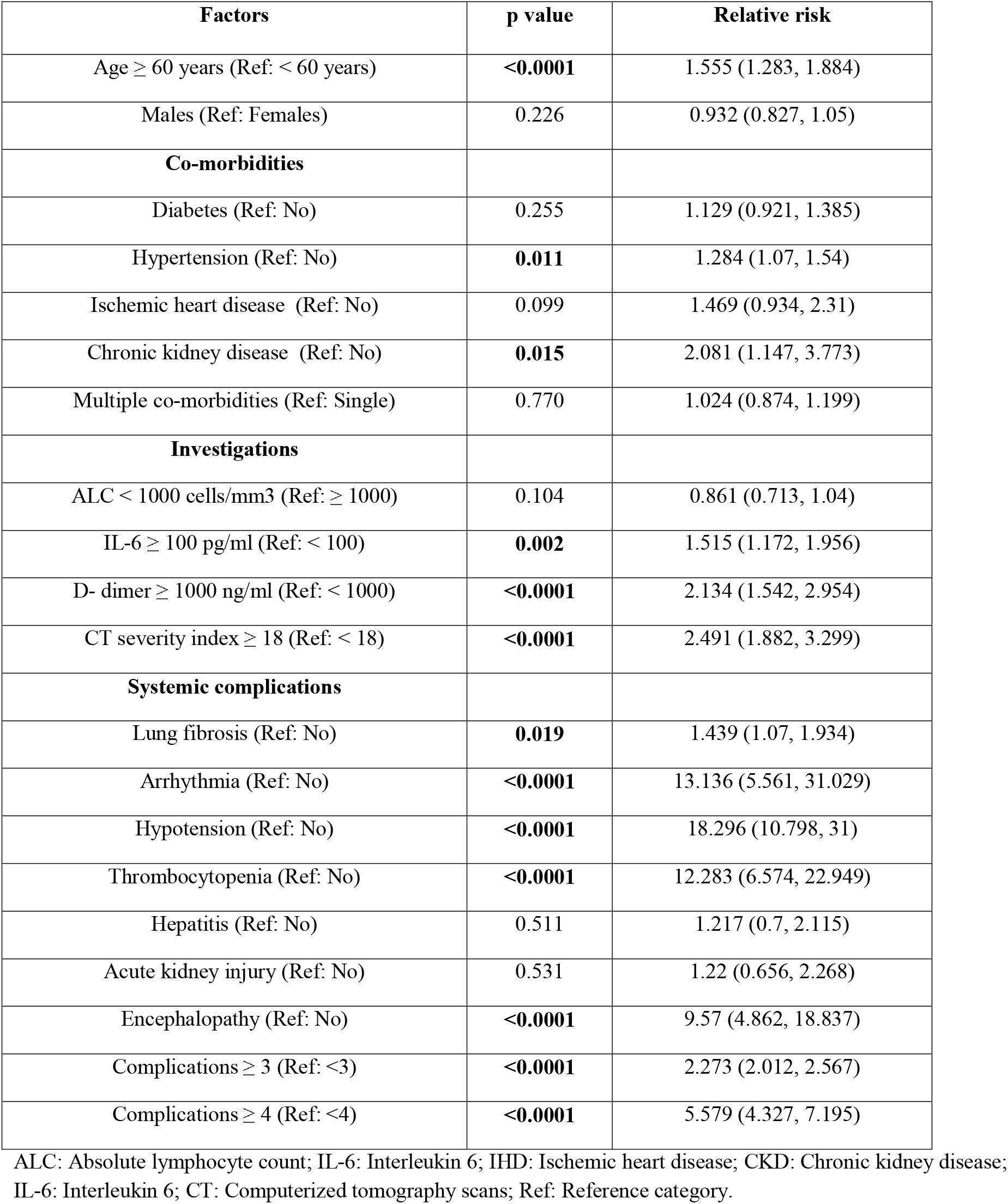
Risk factors associated with death in patients administered Tocilizumab and Steroids.

| Factors | p value | Relative risk |
| --- | --- | --- |
| Age ≥ 60 years (Ref: < 60 years) | **<0.0001** | 1.555 (1.283, 1.884) |
| Males (Ref: Females) | 0.226 | 0.932 (0.827, 1.05) |
| **Co-morbidities** |  |  |
| Diabetes (Ref: No) | 0.255 | 1.129 (0.921, 1.385) |
| Hypertension (Ref: No) | **0.011** | 1.284 (1.07, 1.54) |
| Ischemic heart disease (Ref: No) | 0.099 | 1.469 (0.934, 2.31) |
| Chronic kidney disease (Ref: No) | **0.015** | 2.081 (1.147, 3.773) |
| Multiple co-morbidities (Ref: Single) | 0.770 | 1.024 (0.874, 1.199) |
| **Investigations** |  |  |
| ALC < 1000 cells/mm<sup>3</sup> (Ref: ≥ 1000) | 0.104 | 0.861 (0.713, 1.04) |
| IL-6 ≥ 100 pg/ml (Ref: < 100) | **0.002** | 1.515 (1.172, 1.956) |
| D- dimer ≥ 1000 ng/ml (Ref: < 1000) | **<0.0001** | 2.134 (1.542, 2.954) |
| CT severity index ≥ 18 (Ref: < 18) | **<0.0001** | 2.491 (1.882, 3.299) |
| **Systemic complications** |  |  |
| Lung fibrosis (Ref: No) | **0.019** | 1.439 (1.07, 1.934) |
| Arrhythmia (Ref: No) | **<0.0001** | 13.136 (5.561, 31.029) |
| Hypotension (Ref: No) | **<0.0001** | 18.296 (10.798, 31) |
| Thrombocytopenia (Ref: No) | **<0.0001** | 12.283 (6.574, 22.949) |
| Hepatitis (Ref: No) | 0.511 | 1.217 (0.7, 2.115) |
| Acute kidney injury (Ref: No) | 0.531 | 1.22 (0.656, 2.268) |
| Encephalopathy (Ref: No) | **<0.0001** | 9.57 (4.862, 18.837) |
| Complications ≥ 3 (Ref: <3) | **<0.0001** | 2.273 (2.012, 2.567) |
| Complications ≥ 4 (Ref: <4) | **<0.0001** | 5.579 (4.327, 7.195) |

ALC: Absolute lymphocyte count; IL-6: Interleukin 6; IHD: Ischemic heart disease; CKD: Chronic kidney disease; IL-6: Interleukin 6; CT: Computerized tomography scans; Ref: Reference category.

**Figure 1:**
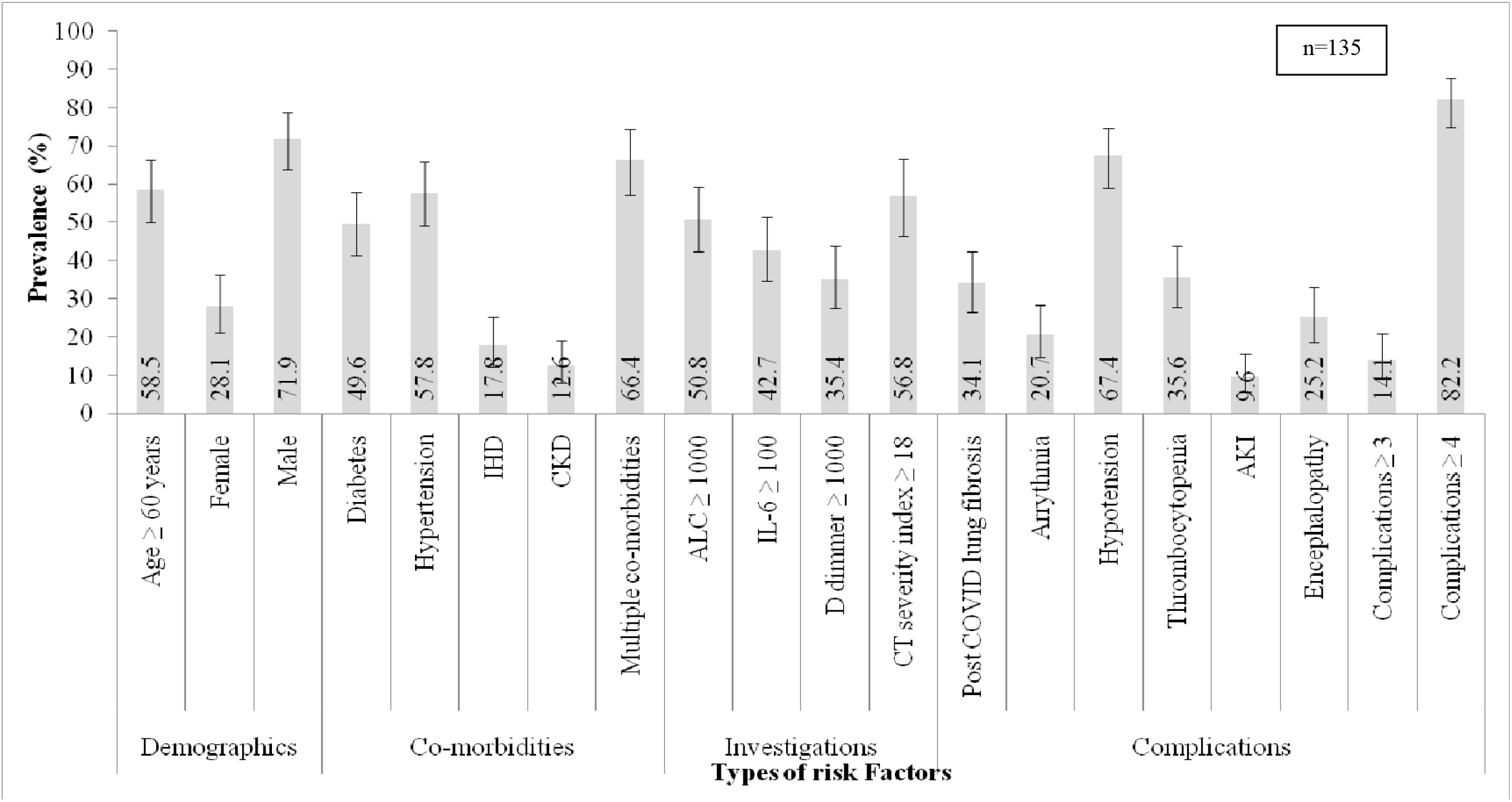
Prevalence of risk factors among COVID-19 patients who died after Tocilizumab and Steroid administration. ALC: Absolute lymphocyte count; IL-6: Interleukin 6; IHD: Ischemic heart disease; CKD: Chronic kidney disease; IL-6: Interleukin 6; CT: Computerized tomography scans.

Out of the 380 patients (73.8 %) who recovered and were discharged from NHRC, 72 (18.9 %) needed short term home oxygen therapy after hospital discharge. Of these, two patients reported worsening of respiratory symptoms at home leading to respiratory failure and death, while 70 patients could be weaned off short term oxygen therapy. Four patients developed late infectious complications (multidermatomal herpes zoster, herpes zoster opthalmicus, acute bacterial cholecystitis and Escherichia coli (E. coli) bacteremia and septic shock) within 1 month of discharge from hospital. Three patients recovered after treatment, but the patient having E. coli bacteremia progressed to septic shock, ARDS and died. One patient developed midbrain encephalitis, 27 days after discharge from hospital and died (Total number of deaths: 139).

## Discussion

The goal of this retrospective observational cohort study conducted at a tertiary level, private hospital in Pune, India was to assess efficacy and safety of combination therapy of TCZ and steroids in treating CRS developing in patients with severe COVID-19. Our cohort consisted of a relatively elderly population (43 % patients ≥ 60 years of age) with pre-existing co-morbidities (72 % having co-morbidities) who developed pneumonia, hyperinflammation and ARDS (100 % patients having increased inflammatory markers, 62.5 % having CT severity score ≥ 15 and 83 % having PaO2/FiO2 < 200 mm Hg). Almost 63 % patients in our cohort required ICU care while 47 % patients needed noninvasive or invasive ventilation to maintain oxygenation. In such a cohort of severely ill patients, administration of TCZ and steroid resulted in clinical improvement in 74 % patients, while 26 % died due to respiratory failure. The response to TCZ was rapid and sustained in majority of patients. Almost 44 % patients could be weaned from ventilator support. However, there was a subset of patients on mechanical ventilation who did show initial improvement after TCZ but subsequently had clinical deterioration and died. This cohort includes 91% of all ICU admissions due to COVID-19 and 96% of all patients receiving mechanical ventilation at NHRC during the said period. To the best of our knowledge, this cohort is the largest reported database of patients who were administered TCZ plus steroids for management of COVID-19 induced CRS and ARDS. Two retrospective cohort studies on TCZ usage from India have been published till date. The strengths of our study compared to the earlier studies include the large number of enrolled patients ^[64]^, strict inclusion and exclusion criteria applied while administering TCZ ^[64,65]^, availability of data about ventilatory outcomes ^[64,65]^, availability of data about systemic complications (including infectious complications) ^[64,65]^ and lower mortality rate ^[65]^.

In our cohort, 37% patients could be managed in isolation wards without need for intensive care. In an ideal scenario, all patients were candidates for ICU care, but overburdened healthcare system and shortage of ICU beds and ventilators meant that they were managed in wards. In addition, 53% patients required only supplemental oxygen prior to recovery and did not progress to mechanical ventilation. As per the CORIMUNO-19 RCT, use of TCZ among patients on supplemental oxygen reduced the need for intensive care, noninvasive or invasive mechanical ventilation by almost 40%. The effect of TCZ was numerically higher if combined with steroids ^[61]^. According to the EMPACTA RCT ^[62]^ and the TESEO cohort ^[32]^, the likelihood of progression to mechanical ventilation was significantly lower among patients who received TCZ plus standard care than among those who received placebo plus standard care. Reduction in need of ICU care can reduce the risk of long-term complications including death and improve health-related quality of life. Preventing progression to mechanical ventilation greatly alters patient outcomes and leads to better utilization of scarce healthcare resources.

Mortality rate in our cohort was 26 %, while mortality rate amongst patients requiring ICU was 40 %. Overall mortality rate was similar to that seen in multiple retrospective cohort studies ^[35, 36, 54, 57, 64]^. Mortality rate amongst patients requiring mechanical ventilation (NIV/IMV) was 56 %. This was higher as compared to other cohort studies ^[40]^. It could be related to delayed intubation and delayed invasive mechanical ventilation policy followed at our hospital. On Relative risk estimation, age ≥ 60 years, presence of multiple co-morbidities, increased baseline inflammatory markers (IL-6 and D-dimer), high CT severity index and development of systemic complications like lung fibrosis, arrhythmia, encephalopathy, thrombocytopenia and hypotension were significant risk factors associated with death. Females had a higher mortality rate than males in our cohort (30.4 % versus 24.9 %, Table 2). Early evidence indicates that males have higher overall burden, but females have a higher relative-risk of COVID-19 mortality in India ^[73]^. Marked sex differences in access to health services, with women being less likely to be admitted to hospital than men might result in more severe cases of COVID-19 among women than men in hospital settings and higher mortality ^[74]^.

TCZ and steroid combination therapy was safe and well tolerated. Infectious complications like confirmed bacterial and fungal infections (including HAP/VAP and bloodstream infections) were seen in 2% patients in the cohort. Low incidence of infectious complications could be because of adequate, prophylactic antibiotic and antifungal therapy prescribed to patients. Meropenem, Teicoplanin and Fluconazole were the commonest antibiotic and antifungal drugs prescribed along with TCZ. Transaminitis or hepatitis, which is an adverse event associated with TCZ, was seen in approximately 10% of patients. However, in view of prescription of multiple drugs like Remdesvir, Fluconazole, Doxycycline, Pirfenidone and Nintedanib along with TCZ, it was difficult to identify the cause of drug induced liver injury. Intestinal perforation after TCZ administration was noted in two patients. Pulmonary (progression of lung fibrosis) and infectious complications were noted even after discharge from hospital. As a result, close follow up of patients for a period of 3 months after hospital discharge is essential for immediate identification of delayed complications.

### Limitations

Our study has several limitations. First, it is not a randomized controlled trial, and therefore unmeasured confounding cannot be ruled out. Second, as for all retrospective studies, some individuals administered TCZ and steroids may be unreported leading to measurement bias and overestimation of safety and efficacy of the combination therapy. Third, a comparator arm was not possible in this pandemic setting. Considering the unavailability of observational or RCT data about efficacy of other regimens to tackle hyperinflammation, worsening of respiratory parameters seen in patients despite receiving potent steroids and life threatening nature of the disease characterized by sudden worsening and rapid progression to respiratory failure over few hours, a comparative arm could not be justified. Fourth, concomitant therapies like antiviral drugs, convalescent plasma, anti-fibrotic agents, supplemental oxygen and ventilation strategies (NIV/IMV) can help in reducing disease severity and clinical improvement. The authors acknowledge the fact that individual contribution of each drug is difficult to estimate. However, considering the predefined criteria for introduction of each drug in patient management, strict inclusion and exclusion criteria applied while administering TCZ and steroids, large number of patients enrolled and improvement in symptoms and oxygenation after TCZ administration, the authors are reasonably confident that this study reliably captures the efficacy of this combination in treating COVID-19 induced CRS. Fifth, an overwhelmed health care system, inadequate workforce and lack of exhaustive reporting could be responsible for underestimation of co-morbidities, presenting symptoms and complications amongst patients in our cohort. Sixth, Body mass index (BMI) could not be calculated for patients who were bed ridden or those requiring mechanical ventilation. Seventh, CT severity index and PaO2/FiO2 ratio was not available for all patients in our cohort. Eighth, Sequential organ failure assessment (SOFA) score was not performed in patients admitted in ICU ^[75]^. Ninth, ventilator parameters like positive end expiratory pressure (PEEP) and plateau pressure were not available for all patients started on NIV or IMV. Tenth, patients were followed up after discharge from hospital for 1 month. As a result long term complications due to immunomodulatory therapy could not be identified.

Despite these limitations, this retrospective cohort study from Western India adds to the growing body of literature on use of TCZ and steroids as an anti-inflammatory combination therapy in treating cytokine storm and resultant ARDS in COVID-19.

## Conclusions

Combination therapy of TCZ and Steroids is likely to be a safe and effective treatment modality in management of COVID-19 associated cytokine release syndrome. Efficacy of this anti-inflammatory combination therapy needs to be validated in large randomized controlled clinical trials.

## Data Availability

De-identified Data is freely available for fellow researchers at link given below

https://figshare.com/s/cebdbaa0b8b84a59ae77

## Abbreviations

CRS: cytokine release syndrome
TCZ: Tocilizumab
NHRC: Noble hospital and Research Centre
COVID-19: coronavirus disease 2019
WHO: World health organization
ARDS: acute respiratory distress syndrome
IL-6: interleukin-6
RCT: randomized controlled trials
RT-PCR: Reverse-transcriptase polymerase chain reaction
HRCT: High resolution computerized tomography scan
CRP: C-reactive protein
IMV: invasive mechanical ventilation
ICMR: Indian council of Medical research
IQR: inter-quartile range
HAP/VAP: Hospital or ventilator associated pneumonia
NRBM: Non re-breathing mask
NIV: noninvasive ventilation
HFNO: High flow nasal oxygen
BIPAP: Bi-level positive airway pressure
GBS: Guillaine Barre syndrome
SGOT/SGPT: serum glutamate oxaloacetate/pyruvate transferase
AKI: Acute kidney injury
CSF: cerebrospinal fluid
MRSA: Methicillin resistant Staphylococcus aureus
E. coli: Escherichia coli
ICU: Intensive care unit
RR: Relative risk
CI: Confidence interval
SOFA: Sequential organ failure assessment
PEEP: positive end expiratory pressure
BMI: Body mass index

## Acknowledgements

Manisha Ghate MD, PhD (National AIDS Research Institute (NARI), Pune, India) - edited the manuscript.

## References

1) Wu Z., McGoogan J.M. Characteristics of and important lessons from the coronavirus disease 2019 (COVID-19) outbreak in China: summary of a report of 72 314 cases from the Chinese Center for Disease Control and Prevention. JAMA. 2020. https://doi.org/10.1001/jama.2020.2648

2) World health organization (WHO): Available at https://www.who.int/emergencies/diseases/novel-coronavirus-2019/technicalguidance/naming-the-coronavirus-disease-(covid-2019)-and-the-virus-that-causes-it.

3) World health organization (WHO) Coronavirus disease (COVID-19) dashboard. Available at: https://covid19.who.int

4) Guan WJ, Ni ZY, Hu Y, Liang WH, Ou CQ, He JX et al; China Medical Treatment Expert Group for Covid-19. Clinical Characteristics of Coronavirus Disease 2019 in China. N Engl J Med. 2020 Apr 30; 382(18): 1708–1720. https://doi.org/10.1056/NEJMoa2002032

5) Chen N, Zhou M, Dong X, Qu J, Gong F, Han Y et al. Epidemiological and clinical characteristics of 99 cases of 2019 novel coronavirus pneumonia in Wuhan, China: a descriptive study. Lancet. 2020 Feb 15; 395(10223):507–513. https://doi.org/10.1016/S0140-6736(20)30211-7

6) Wang D, Hu B, Hu C, Zhu F, Liu X, Zhang J et al. Clinical Characteristics of 138 Hospitalized Patients With 2019 Novel Coronavirus-Infected Pneumonia in Wuhan, China. JAMA. 2020 Mar 17; 323(11):1061–1069. https://doi.org/10.1001/jama.2020.1585

7) Yang X., Yu Y., Xu J., Shu H., Xia J., Liu H. Clinical course and outcomes of critically ill patients with SARS-CoV-2 pneumonia in Wuhan, China: a single-centered, retrospective, observational study. Lancet Respir Med. 2020 https://doi.org/10.1016/S2213-2600(20)30079-5

8) Bhatraju PK, Ghassemieh BJ, Nichols M, Kim R, Jerome KR, Nalla AK, et al. Covid-19 in critically ill patients in the Seattle region. N Engl J Med. 2020; 382 (21):2012–2022. https://doi.org/10.1056/NEJMoa2004500

9) Yang X, Yu Y, Xu J, Shu H, Xia J, Liu H, et al. Clinical course and outcomes of critically ill patients with SARS-CoV-2 pneumonia in Wuhan, China. Lancet Respir Med. 2020; 8(5): 475–481. https://doi.org/10.1016/S2213-2600(20)30079-5

10) Gupta S, Hayek SS, Wang W, Chan L, Mathews KS, Melamed ML, et al; STOP-COVID Investigators. Factors associated with death in critically ill patients with coronavirus disease 2019 in the US. Published online July 15, 2020. JAMA Intern Med. https://doi.org/10.1001/jamainternmed.2020.3596

11) Pedersen SF, Ho YC. SARS-CoV-2: a storm is raging. J Clin Invest. 2020 May 1; 130(5): 2202–2205. https://doi.org/10.1172/JCI137647

12) García LF. Immune Response, Inflammation, and the Clinical Spectrum of COVID-19. Front Immunol. 2020 Jun 16; 11: 1441. https://doi.org/10.3389/fimmu.2020.01441

13) Catanzaro M, Fagiani F, Racchi M, Corsini E, Govoni S, Lanni C. Immune response in COVID-19: addressing a pharmacological challenge by targeting pathways triggered by SARS-CoV-2. Signal Transduct Target Ther. 2020 May 29; 5(1):84. https://doi.org/10.1038/s41392-020-0191-1

14) Shi Y, Wang Y, Shao C, Huang J, Gan J, Huang X, et al. COVID-19 infection: the perspectives on immune responses. Cell Death Differ. 2020 May; 27(5): 1451–1454. https://doi.org/10.1038/s41418-020-0530-3

15) Chen G, Wu D, Guo W, Cao Y, Huang D, Wang H et al. Clinical and immunological features of severe and moderate coronavirus disease 2019. J Clin Invest. 2020 May 1; 130(5): 2620–2629. https://doi.org/10.1172/JCI137244

16) Wu C, Chen X, Cai Y, Xia J, Zhou X, Xu S, et al. Risk factors associated with acute respiratory distress syndrome and death in patients with coronavirus disease 2019 pneumonia in Wuhan, China. JAMA Intern Med. (2020). https://doi.org/10.1001/jamainternmed.2020.0994

17) Herold T, Jurinovic V, Arnreich C, Lipworth BJ, Hellmuth JC, von Bergwelt-Baildon M, et al. Elevated levels of IL-6 and CRP predict the need for mechanical ventilation in COVID-19. J Allergy Clin Immunol 2020; 146(1): 128-136.e4.

18) Del Valle DM, Kim-Schulze S, Huang HH, Beckmann ND, Nirenberg S, Wang B, et al. An inflammatory cytokine signature predicts COVID-19 severity and survival. Nat Med. 2020 Oct; 26(10): 1636–1643. https://doi.org/10.1038/s41591-020-1051-9

19) McGonagle D, Sharif K, O’Regan A, Bridgewood C. The Role of Cytokines including Interleukin-6 in COVID-19 induced Pneumonia and Macrophage Activation Syndrome-Like Disease. Autoimmun Rev. 2020 Jun; 19(6):102537. https://doi.org/10.1016/j.autrev.2020.102537

20) WHO Rapid Evidence Appraisal for COVID-19 Therapies (REACT) Working Group, Sterne JAC, Murthy S, Diaz JV, Slutsky AS, Villar J, Angus DC et al. Association Between Administration of Systemic Corticosteroids and Mortality Among Critically Ill Patients With COVID-19: A Meta-analysis. JAMA. 2020 Oct 6; 324(13):1330–1341. https://doi.org/10.1001/jama.2020.17023

21) RECOVERY Collaborative Group, Horby P, Lim WS, Emberson JR, Mafham M, Bell JL, Linsell L et al. Dexamethasone in Hospitalized Patients with Covid-19 - Preliminary Report. N Engl J Med. 2020 Jul 17: EJMoa2021436. https://doi.org/10.1056/NEJMoa2021436

22) Fadel R, Morrison AR, Vahia A, Smith ZR, Chaudhry Z, Bhargava P et al ; Henry Ford COVID-19 Management Task Force. Early Short-Course Corticosteroids in Hospitalized Patients with COVID-19. Clin Infect Dis. 2020 Nov 19; 71(16):2114–2120. https://doi.org/10.1093/cid/ciaa601

23) Nelson BC, Laracy J, Shoucri S, Dietz D, Zucker J, Patel N et al. Clinical Outcomes Associated with Methylprednisolone in Mechanically Ventilated Patients with COVID-19. Clin Infect Dis. 2020 Aug 9:ciaa1163. https://doi.org/10.1093/cid/ciaa1163

24) Zhang C, Wu Z, Li JW, Zhao H, Wang GQ. Cytokine release syndrome in severe COVID-19: interleukin-6 receptor antagonist tocilizumab may be the key to reduce mortality. Int J Antimicrob Agents. 2020; 55(5): 105954. https://doi.org/10.1016/j.ijantimicag.2020.105954

25) Actemra (tocilizumab) Prescribing information. South San Francisco, CA: Genentech, Inc; 2019.

26) Navarro G., Taroumian S., Barroso N., Duan L., Furst D. Tocilizumab in rheumatoid arthritis: a meta-analysis of efficacy and selected clinical conundrums. Semin Arthritis Rheum. 2014; 43: 458–469.

27) Yokota S., Miyamae T., Imagawa T., Iwata N., Katakura S., Mori M. Therapeutic efficacy of humanized recombinant anti-interleukin-6 receptor antibody in children with systemic-onset juvenile idiopathic arthritis. Arthritis Rheum. 2005; 52: 818–825.

28) Nishimoto N., Kanakura Y., Aozasa K., Johkoh T., Nakamura M., Nakano S. Humanized anti-interleukin-6 receptor antibody treatment of multicentric Castleman disease. Blood. 2005; 106:2627–2632.

29) Ito H., Takazoe M., Fukuda Y., Hibi T., Kusugami K., Andoh A. A pilot randomized trial of a human anti-interleukin-6 receptor monoclonal antibody in active Crohn’s disease. Gastroenterology. 2004; 126: 989–996.

30) Le R.Q., Li L., Yuan W., Shord S.S., Nie L., Habtemariam B.A. FDA approval summary: tocilizumab for treatment of chimeric antigen receptor T cell-induced severe or life-threatening cytokine release syndrome. Oncologist. 2018; 23: 943–947.

31) CLINICAL MANAGEMENT PROTOCOL: COVID-19. Government of India Ministry of Health and Family Welfare Directorate General of Health Services (EMR Division). Version 5. Available at : https://www.mohfw.gov.in/pdf/UpdatedClinicalManagementProtocolforCOVID19dated03072020.pdf

32) Guaraldi G, Meschiari M, Cozzi-Lepri A, Milic J, Tonelli R, Menozzi M, et al. Tocilizumab in patients with severe COVID-19: a retrospective cohort study. Lancet Rheumatol. 2020 Aug; 2(8): e474–e484. https://doi.org/10.1016/S2665-9913(20)30173-9

33) Kewan T, Covut F, Al-Jaghbeer MJ, Rose L, Gopalakrishna KV, Akbik B. Tocilizumab for treatment of patients with severe COVID-19: A retrospective cohort study. E Clinical Medicine. 2020 Jun 20; 24:100418. https://doi.org/10.1016/j.eclinm.2020.100418

34) Xu X, Han M, Li T, Sun W, Wang D, Fu B, et al. Effective treatment of severe COVID-19 patients with tocilizumab. Proc Natl Acad Sci USA 2020; 117: 10970–10975. https://doi.org/10.1073/pnas.2005615117

35) Zain Mushtaq M, Bin Zafar Mahmood S, Jamil B, Aziz A, Ali SA. Outcome of COVID-19 patients with use of Tocilizumab: A single center experience. Int Immunopharmacol. 2020 Nov; 88:106926. https://doi.org/10.1016/j.intimp.2020.106926

36) Luo P, Liu Y, Qiu L, Liu X, Liu D, Li J. Tocilizumab treatment in COVID-19: A single center experience. J Med Virol. 2020 Jul; 92(7): 814–818. https://doi.org/10.1002/jmv.25801

37) Campins L, Boixeda R, Perez-Cordon L, Aranega R, Lopera C, Force L. Early tocilizumab treatment could improve survival among COVID-19 patients. Clin Exp Rheumatol 2020; 38: 578.

38) Rossotti R, Travi G, Ughi N, Corradin M, Baiguera C, Fumagalli R, et al. Safety and efficacy of anti-IL6-receptor tocilizumab use in severe and critical patients affected by coronavirus disease 2019: a comparative analysis. J Infect 2020; 81(4): e11–e17. https://doi.org/10.1016/j.jinf.2020.07.008

39) Biran N, Ip A, Ahn J, Go RC, Wang S, Mathura S et al. Tocilizumab among patients with COVID-19 in the intensive care unit: a multicentre observational study. Lancet Rheumatol. 2020 Oct; 2(10): e603–e612. https://doi.org/10.1016/S2665-9913(20)30277-0

40) Somers EC, Eschenauer GA, Troost JP, Golob JL, Gandhi TN, Wang L, et al. Tocilizumab for treatment of mechanically ventilated patients with COVID-19. Clin Infect Dis. 2020 Jul 11:ciaa954. https://doi.org/10.1093/cid/ciaa954

41) Antony SJ, Davis MA, Davis MG, Almaghlouth NK, Guevara R, Omar F, et al. Early use of tocilizumab in the prevention of adult respiratory failure in SARS-CoV-2 infections and the utilization of interleukin-6 levels in the management. J Med Virol. 2020 Jul 9:10.1002/jmv.26288. https://doi.org/10.1002/jmv.26288

42) Sanz Herrero F, Puchades Gimeno F,Ortega Garcia P, Ferrer Gomez C, Ocete Mochon MD, Garcia Deltoro M. Methylprednisolone added to tocilizumab reduces mortality in SARS-CoV-2 pneumonia: an observational study. J Intern Med 2020 June 30.

43) Jordan SC, Zakowski P, Tran HP, Smith EA, Gaultier C, Marks G, et al. Compassionate use of tocilizumab for treatment of SARS-CoV-2 pneumonia. Clin Infect Dis. 2020 Jun 23:ciaa812. https://doi.org/10.1093/cid/ciaa812

44) Quartuccio L, Sonaglia A, McGonagle D, Fabris M, Peghin M, Pecori D, et al. Profiling COVID-19 pneumonia progressing into the cytokine storm syndrome: results from a single Italian Centre study on tocilizumab versus standard of care. J Clin Virol. 2020 Aug; 129: 104444. https://doi.org/10.1016/j.jcv.2020.104444

45) Rojas-Marte G, Khalid M, Mukhtar O, Hashmi AT, Waheed MA, Ehrlich S, et al. Outcomes in patients with severe COVID-19 disease treated with tocilizumab: a case-controlled study. QJM. 2020 Aug 1; 113 (8): 546–550. https://doi.org/10.1093/qjmed/hcaa206

46) Price CC, Altice FL, Shyr Y, Koff A, Pischel L, Goshua G, et al. Tocilizumab treatment for cytokine release syndrome in hospitalized COVID-19 patients: survival and clinical outcomes. Chest. 2020 Oct; 158(4): 1397–1408. https://doi.org/10.1016/j.chest.2020.06.006

47) Pérez-Sáez MJ, Blasco M, Redondo-Pachón D, Ventura-Aguiar P, Bada-Bosch T, Pérez-Flores I, et al. Use of tocilizumab in kidney transplant recipients with COVID-19. Am J Transplant. 2020 Nov; 20(11):3182–3190. https://doi.org/10.1111/ajt.16192

48) Knorr JP, Colomy V, Mauriello CM, Ha S. Tocilizumab in patients with severe COVID-19: a single-center observational analysis. J Med Virol. 2020 Nov; 92(11):2813–2820. https://doi.org/10.1002/jmv.26191

49) Campochiaro C, Della-Torre E, Cavalli G, De Luca G, Ripa M, Boffini N, et al. Efficacy and safety of tocilizumab in severe COVID-19 patients: a single centre retrospective cohort study. Eur J Intern Med. 2020 Jun; 76: 43–49. https://doi.org/10.1016/j.ejim.2020.05.021

50) Mikulska M, Nicolini LA, Signori A, Di Biagio A, Sepulcri C, Russo C et al. Tocilizumab and steroid treatment in patients with COVID-19 pneumonia. PLoS One. 2020 Aug 20; 15(8):e0237831. https://doi.org/10.1371/journal.pone.0237831

51) Colaneri M, Bogliolo L, Valsecchi P,et al. Tocilizumab for treatment of severe COVID-19 patients: preliminary results from SMAtteo COvid19 Registry (SMACORE). Microorganisms 2020; 8: 695.

52) Klopfenstein T, Zayet S, Lohse A, Balblanc JC, Badie J, Royer PY, et al. Tocilizumab therapy reduced intensive care unit admissions and/or mortality in COVID-19 patients. Med Mal Infect 2020; 50: 397–400. https://doi.org/10.1016/j.medmal.2020.05.001

53) Ip A, Berry DA, Hansen E, Goy AH, Pecora AL, Sinclaire BA et al. Hydroxychloroquine and tocilizumab therapy in COVID-19 patients-An observational study. PLoS One. 2020 Aug 13; 15(8):e0237693. https://doi.org/10.1371/journal.pone.0237693

54) Toniati P, Piva S, Cattalini M, Garrafa E, Regola F, Castelli F, et al. Tocilizumab for the treatment of severe COVID-19 pneumonia with hyperinflammatory syndrome and acute respiratory failure: a single center study of 100 patients in Brescia, Italy. Autoimmun Rev 2020; 19: 102568.

55) Alattar R, Ibrahim TBH, Shaar SH, Abdalla S, Shukri K, Daghfal JN, et al. Tocilizumab for the treatment of severe coronavirus disease 2019. J Med Virol. 2020 Oct; 92(10):2042–2049. https://doi.org/10.1002/jmv.25964

56) Potere N, Di Nisio M, Cibelli D, Scurti R, Frattari A, Porreca E, et al. Interleukin-6 receptor blockade with subcutaneous tocilizumab in severe COVID-19 pneumonia and hyperinflammation: a case-control study. Ann Rheum Dis. 2020 Jul 9: annrheumdis-2020–218243. https://doi.org/10.1136/annrheumdis-2020-218243

57) Gupta S, Wang W, Hayek SS, Chan L, Mathews KS, Melamed ML et al; STOP-COVID Investigators. Association between Early Treatment with Tocilizumab and Mortality among Critically Ill Patients with COVID-19. JAMA Intern Med. 2020 Oct 20:e206252. https://doi.org/10.1001/jamainternmed.2020.6252

58) Capra R, De Rossi N, Mattioli F, Romanelli G, Scarpazza C, Sormani MP et al. Impact of low dose tocilizumab on mortality rate in patients with COVID-19 related pneumonia. Eur J Intern Med. 2020 Jun; 76: 31–35. https://doi.org/10.1016/j.ejim.2020.05.009

59) Salvarani C, Dolci G, Massari M, Merlo DF, Cavuto S, Savoldi L et al; RCT-TCZ-COVID-19 Study Group. Effect of Tocilizumab vs Standard Care on Clinical Worsening in Patients Hospitalized with COVID-19 Pneumonia: A Randomized Clinical Trial. JAMA Intern Med. 2020 Oct 20:e206615. https://doi.org/10.1001/jamainternmed.2020.6615

60) Stone JH, Frigault MJ, Serling-Boyd NJ, Fernandes AD, Harvey L, Foulkes AS et al; BACC Bay Tocilizumab Trial Investigators. Efficacy of Tocilizumab in Patients Hospitalized with Covid-19. N Engl J Med. 2020 Oct 21:EJMoa2028836. https://doi.org/10.1056/NEJMoa2028836

61) Hermine O, Mariette X, Tharaux PL, Resche-Rigon M, Porcher R, Ravaud P; CORIMUNO-19 Collaborative Group. Effect of Tocilizumab vs Usual Care in Adults Hospitalized With COVID-19 and Moderate or Severe Pneumonia: A Randomized Clinical Trial. JAMA Intern Med. 2020 Oct 20:e206820. https://doi.org/10.1001/jamainternmed.2020.6820

62) Salama C, Han J, Yau L, Reiss WG, Kramer B, Neidhart JD et al. Tocilizumab in Patients Hospitalized with Covid-19 Pneumonia. N Engl J Med. 2020 Dec 17. https://doi.org/10.1056/NEJMoa2030340

63) Rosas I, Bräu N, Waters M, et al. Tocilizumab in hospitalized patients with COVID-19 pneumonia. medRxiv. https://doi.org/10.1101/2020.08.27.20183442

64) Patel A, Shah K, Dharsandiya M, Patel K, Patel T, Patel M, Reljic T, Kumar A. Safety and efficacy of tocilizumab in the treatment of severe acute respiratory syndrome coronavirus-2 pneumonia: A retrospective cohort study. Indian J Med Microbiol. 2020 Jan-Mar; 38(1):117–123. https://doi.org/10.4103/ijmm.IJMM_20_298

65) Gokhale Y, Mehta R, Karnik N, Kulkarni U, Gokhale S. Tocilizumab improves survival in patients with persistent hypoxia in severe COVID-19 pneumonia. E Clinical Medicine. 2020; 24:100467. https://doi.org/10.1016/j.eclinm.2020.100467

66) Francone M, Iafrate F, Masci GM, Coco S, Cilia F, Manganaro Let al. Chest CT score in COVID-19 patients: correlation with disease severity and short-term prognosis. Eur Radiol. 2020 Dec; 30(12): 6808–6817. https://doi.org/10.1007/s00330-020-07033-y

67) Elavarasi A, Prasad M, Seth T, Sahoo RK, Madan K, Nischal N, Soneja M, Sharma A, Maulik SK, Shalimar Garg P. Chloroquine and Hydroxychloroquine for the Treatment of COVID-19: a Systematic Review and Meta-analysis. J Gen Intern Med. 2020 Nov; 35(11):3308–3314. https://doi.org/10.1007/s11606-020-06146-w

68) Cao B, Wang Y, Wen D, Liu W, Wang J, Fan G et al. A Trial of Lopinavir-Ritonavir in Adults Hospitalized with Severe Covid-19. N Engl J Med. 2020 May 7; 382(19):1787–1799. https://doi.org/10.1056/NEJMoa2001282

69) RECOVERY Collaborative Group. Lopinavir-ritonavir in patients admitted to hospital with COVID-19 (RECOVERY): a randomised, controlled, open-label, platform trial. Lancet. 2020 Oct 5; 396(10259):1345–1352. https://doi.org/10.1016/S0140-6736(20)32013-4

70) Beigel JH, Tomashek KM, Dodd LE, Mehta AK, Zingman BS, Kalil AC et al; ACTT-1 Study Group Members. Remdesivir for the Treatment of Covid-19 - Final Report. N Engl J Med. 2020 Nov 5; 383(19):1813–1826. https://doi.org/10.1056/NEJMoa2007764

71) Acute kidney injury (AKI) guidelines. Available at : https://kdigo.org/wp-content/uploads/2016/10/KDIGO-2012-AKI-Guideline-English.pdf

72) Ellul MA, Benjamin L, Singh B, Lant S, Michael BD, Easton A et al. Neurological associations of COVID-19. Lancet Neurol. 2020 Sep; 19(9): 767–783. https://doi.org/10.1016/S1474-4422(20)30221-0

73) Joe W, Kumar A, Rajpal S, Mishra U.S., Subramanian SV. Equal risk, unequal burden? Gender differentials in COVID-19 mortality in India. J Glob Health Sci. 2020 Jun;2(1):e17

74) Dehingia N, Raj A. Sex differences in COVID-19 case fatality: do we know enough? Lancet Glob Health. 2021 Jan; 9(1): e14–e15. https://doi.org/10.1016/S2214-109X(20)30464-2

75) Jones AE, Trzeciak S, Kline JA. The Sequential Organ Failure Assessment score for predicting outcome in patients with severe sepsis and evidence of hypoperfusion at the time of emergency department presentation. Crit Care Med. 2009; 37(5):1649–1654. https://doi.org/10.1097/CCM.0b013e31819def97

